# Multiplexed and extraction-free amplification for simplified SARS-CoV-2 RT-PCR tests

**DOI:** 10.1101/2020.05.21.20106195

**Authors:** Samantha A. Byrnes, Ryan Gallagher, Amy Steadman, Crissa Bennett, Rafael Rivera, Corrie Ortega, S. Timothy Motley, Paras Jain, Bernhard H. Weigl, John T. Connelly

**Author notes:** Address for correspondence: Intellectual Ventures Laboratory, 14360 SE Eastgate Way Bellevue, 98007, WA, USA, Samantha A. Byrnes, Ryan Gallagher, John T. Connelly.

## Abstract

The rapid onset of the global COVID-19 pandemic has led to multiple challenges for accurately diagnosing the infection. One of the main bottlenecks for COVID-19 detection is reagent and material shortages for sample collection, preservation, and purification prior to testing. Currently, most authorized diagnostic tests require RNA extraction from patient samples and detection by reverse transcription polymerase chain reaction (RT-PCR). However, RNA purification is expensive, time consuming, and requires technical expertise to perform. Additionally, there have been reported shortages of the RNA purification kits needed for most tests. With these challenges in mind, we report on extraction-free amplification of SARS-CoV-2 RNA directly from patient samples. In addition, we have developed a multiplex RT-PCR using the CDC singleplex targets. This multiplex has a limit of detection of 2 copies/μL. We have demonstrated these improvements to the current diagnostic workflow, which reduce complexity and cost, minimize reagent usage, expedite time to results, and increase testing capacity.

## Introduction

In December 2019 a pneumonia of unknown cause was detected in Wuhan, China and reported to the WHO(1). The novel coronavirus disease (COVID-19) spread rapidly on a global scale. At the time of this writing, there are more than four and a half million cases of COVID-19 and over 300,000 deaths reported to the WHO with numbers increasing daily(2). In January, severe acute respiratory syndrome coronavirus 2 (SARS-CoV-2) was confirmed as the infectious agent and the full genome sequence(3) was published shortly after, enabling the development of critical diagnostic tools. Testing is at the core of the COVID-19 response and is critical to abating this global pandemic.

Currently, the vast majority of approved diagnostics require RNA extraction from respiratory tract specimens and detection by reverse transcription polymerase chain reaction (RT-PCR)(4). RNA extraction reproducibly isolates and purifies RNA from upper respiratory tract samples, including nasopharyngeal (NP) swabs stored in Viral/Universal Transport Media (VTM/UTM), with minimal loss of original sample. However, RNA purification kits are expensive and sample processing adds complexity and time to the diagnostic workflow. Most concerning, months into the COVID-19 pandemic, testing remains dangerously inadequate in part due to shortages of test materials, including specimen swabs, transport media, and extraction reagents. In addition, many approved diagnostics, such as the CDC 2019-nCoV Real-Time RT-PCR Diagnostic Panel, require running three singleplex (one target at a time) reactions per sample(5), limiting testing throughput.

Streamlining the RT-PCR diagnostic workflow to reduce complexity, time, and reagent usage could increase testing consistency and capacity. Removing RNA extraction and identifying alternative reagents that demonstrate equivalent performance to those used in approved COVID-19 diagnostics can increase reagent options and alleviate known bottlenecks. Efforts are underway by multiple groups to achieve these goals. Amplification of SARS-CoV-2 direct from patient swabs in simple buffers such as PBS(6) or TE (Tris, EDTA)(7) has been recently demonstrated. Although promising, these workflows do not account for the current clinical practices of collecting swabs in transport media. Ideally, simplification of COVID-19 diagnostic tests should align with current clinical practices. Beltran-Pavez *et al* demonstrated direct amplification from transport media, but only tested a single type of VTM which resulted in significant amplification inhibition(8). With current reagent shortages, improvements to diagnostic test workflow should explore a variety of transport media, including options that can easily be made in-house. The CDC has an authorized standard operating procedure (SOP) for preparing VTM (SOP#: DSR-052-02. 2020) from readily available reagents when commercial sources of VTM are unavailable(9).

Given the current landscape of SARS-CoV-2 diagnostics and the need to increase testing capacity, we have explored directly amplifying viral RNA from multiple commercial transport media in combination with testing five RT-PCR master mixes (MMs) from different manufactures. Selection criteria were based on compatibility with specimen collection types, such as NP swabs, and RT-PCR formulations marketed as having increased inhibitor tolerance and reported to support direct RT-PCR amplification(10–12). Four of the five MMs are authorized for use with the CDC 2019-nCoV Real-Time RT-PCR Diagnostic Panel(5), and one has shown encouraging results for direct RT-PCR amplification from clinical samples(13).

In addition to exploring extraction-free amplification directly from clinical samples, we also tested the CDC 2019-nCoV Real-Time RT-PCR Diagnostic Panel as a multiplex RT-PCR compared to the three singleplex reactions required currently(14). The CDC 2019-nCoV Real-Time RT-PCR Diagnostic Panel (referred to as the “CDC singleplex test” from here on) was released in early February to all US states and 30 international locations. Over 50 molecular COVID-19 diagnostics have received FDA emergency use authorization (EUA) since the CDC singleplex test was implemented(15). Multiplexing of the CDC targets has been demonstrated(16) and five commercial COVID-19 molecular diagnostics are multiplex derivatives of the CDC singleplex test. However, four of the five multiplex tests require specialized equipment, such as the BD MAX system(17), and one of the five is approved with a single RT-PCR MM(18) compared to the four MMs approved with the CDC singleplex test, limiting reagent options. Demonstrating performance of an extraction-free multiplex RT-PCR from VTM samples for SARS-CoV-2 detection would reduce complexity, time, and reagent bottlenecks while enhancing throughput and remaining in line with current clinical practices.

With these goals in mind, we demonstrate that direct amplification from multiple transport media when paired with inhibitor-tolerant MM formulations is equivalent to approved, more complex diagnostic protocols. In addition, we have developed a multiplex RT-PCR using the CDC singleplex targets. This multiplex has a limit of detection of 2 copies/μL (c/μL) of patient sample. We have successfully demonstrated improvements to the current diagnostic workflow, which reduce complexity, may alleviate reagent shortages, shorten time to results, and increase throughput. The improvements and findings described here may also be suitable for other pathogen-detection applications and diagnostics.

## Materials and Methods

### Reagents

All primers and probes, purified 2019-nCoV_N RNA, and Hs_RPP30 human RNA were purchased from IDT (Coralville, IA, USA). The Research Use Only (RUO) QIAamp Viral Mini Kit for RNA extraction was purchased from Qiagen (Hilden, Germany). We choose the RUO kit to avoid exacerbating shortages of the clinical versions. The GoTaq 1-step RT-qPCR kit was purchased from Promega (Madison, WI, USA). The TaqPath 1-step RT-qPCR Master Mix CG, Remel M4RT viral transport media (VTM), Hanks’ Balanced Salt Solution, heat-inactivated fetal bovine serum, and gentamicin were purchased were purchased from ThermoFisher Scientific (Waltham, MA, USA). The Luna Universal Probe One Step RT qPCR kit was purchased from NEB (Ipswich, MA, USA). The qScript XLT 1-Step RT-qPCR ToughMix and UltraPlex 1-Step ToughMix were purchased from QuantaBio (Beverly, MA, USA). Molecular biology grade water was purchased from Fisher Scientific (Waltham, MA, USA). The S2 VTM was purchased from S2M Enterprises (Spokane Valley, WA, USA). Amphotericin B and absolute ethanol were purchased from Sigma-Aldrich (St. Louis, MO, USA).

The 10 mM Tris, pH 8, was prepared in-house with molecular biology grade water. The in-house VTM (GG-VTM) was prepared according SOP# DSR-052-01 from the USA Centers for Disease Control and Prevention(9).

### Clinical samples

We received 90 de-identified samples from the Washington State Department of Health (DOH) Public Health Laboratories (Shoreline, WA, USA). The samples were collected in March 2020 from nasopharyngeal (NP) swabs added to VTM and included 30 SARS-CoV-2 positives and 60 negatives. Matched purified RNA was provided for all positive samples. Half of the negative samples were also provided as purified RNA, but we did not receive the matched VTM. The remaining 30 negative samples were provided as aliquots of VTM from NP samples and were purified prior to analysis. Viral loads for all samples were determined using the CDC singleplex test. All samples were discarded and de-identified and therefore did not require IRB approval for use.

We also received 78 clinical samples from Discovery Life Sciences (Los Osos, CA, USA), which were negative for SARS-CoV-2, but 17 were positive for a variety of other respiratory infections including: influenza A and B, respiratory syncytial virus, pneumoniae, and coronavirus NL63, HKU1, or OC43. These samples arrived as NP swabs in VTM and RNA was purified from each sample prior to testing in the SARS-CoV-2 RT-PCR.

### RNA purification from clinical samples

For RNA extraction, 140 μL of sample was purified using the QIAamp Viral Mini Kit according to the manufacturer’s protocol(19). Briefly, AVL lysis buffer was added to the sample and incubated at room temperature for 10 minutes. After incubation, molecular grade absolute ethanol was added and the solution was transferred to a QIAamp mini spin column. The column was centrifuged for 1 minute at 6000 x g and the flow through was discarded. The spin column was washed with AW1 buffer and centrifuged for 1 minute at 6000 x g. A second wash was done using AW2 buffer and centrifuged for 3 minutes at 20000 x g and flow through was discarded. Finally, the purified RNA was eluted in 140 μL of provided elution buffer according to CDC recommendations(14). Purified RNA was stored at -80 °C. Positive and negative extraction controls were made by spiking either human and SARS-CoV-2 RNA or just human RNA into the elution buffer and extracting alongside the clinical samples.

### In silico analysis

All CDC primers and probes sequences were analyzed in Geneious Prime version 2020.0.3 (https://www.geneious.com) and screened for unfavorable folding and oligomer interactions using AutoDimer Version 1.0(20) with the following conservative parameters: Minimum SCORE Requirement: 3; Na+ 0.085 M; Temp for dG calc 37°C; Total Strand Conc 1.0 μM.

### Singleplex and multiplex RT-PCR for SARS-CoV-2

All RT-PCR reactions were run using the BioRad CFX96 Real-Time PCR Detection System (Hercules, CA, USA) with fluorescent data collected during the annealing step. The final primers and probe concentrations for the singleplex (one target at a time) reactions were 500 nM and 125 nM, respectively(14). The final primers and probes concentrations for the multiplex (all three targets simultaneously) reaction were 500 nM and 250 nM, respectively. All primer and probe sequences are available in **Table S1**. For each reaction, 5 μL of sample was added to 15 μL of amplification mix. The protocols used for each MM are outlined in **Table 1**.

**Table 1.** Overview of RT-PCR protocols for each MM tested. All reactions included 45 cycles of denaturation and annealing based on the recommendation from the CDC for the SARS-CoV-2 N1 and N2 targets(14). The protocols were the same for both the singleplex and multiplex reactions.

| Master Mix | Initial Step | Reserve Transcriptase | Initial Denaturation | Thermal Cycling (45x) |
| --- | --- | --- | --- | --- |
| TaqPath, ThermoFisher | 25 °C, 2 min | 50 °C, 15 min | 95 °C, 2 min | 95 °C 3 sec, 55 °C 30 sec |
| GoTaq, Promega | - | 45 °C, 15 min | 95 °C, 2 min | 95 °C 15 sec, 55 °C 60 sec |
| UltraPlex, QuantaBio | - | 50 °C, 10 min | 95 °C, 3 min | 95 °C 3 sec, 55 °C 30 sec |
| qScript, QuantaBio | - | 50 °C, 10 min | 95 °C, 1 min | 95 °C 3 sec, 55 °C 30 sec |
| LunaScript, NEB | - | 50 °C, 10 min | 95 °C, 1 min | 95 °C 10 sec, 55 °C 30 sec |

### Multiplex LoD

Multiplex RT-PCR was performed using TaqPath MM with 10-fold dilutions of RNA (2×10^1^ – 2×10^5^ each of human RNA and SARS-CoV-2 RNA per reaction) and compared to corresponding CDC singleplex test for each target (N1, N2, and RP). N3 primers and probe were omitted as they were removed from the CDC test prior to the onset of this work. Additionally, multiplex reactions were tested with 0.8 – 8.0 c/μL (c/μL) SARS-CoV-2 RNA with increasing amounts of human RNA input (10^1^ – 10^4^ c/μL) to assess competitive inhibition. Purified SARS-CoV-2 RNA was quantified in-house using the BioRad QX200 Digital Droplet PCR System (Hercules, CA, USA).

### Singleplex v. multiplex amplification with patient samples

Purified RNA from clinical patient samples provided the by the Washington State Department of Health (DOH) Public Health Laboratories was used as input in both singleplex and multiplex RT-PCR. For each reaction, 5 μL of sample was added to 15 μL of amplification mix, as described above.

### Impact of viral transport media on RT-PCR

The impact of three VTMs (S2, M4RT, and GG-VTM) on five different RT-PCR MMs was evaluated by spiking SARS-CoV-2 and human RNA into stock and diluted VTM. Each of the VTMs was diluted with 10 mM Tris, pH 8, at ratios of 1:10, 1:100, or 1:1000. RNA was spiked in after dilution to maintain a consistent concentration across dilutions. Control samples were prepared by spiking RNA into 10 mM Tris, pH 8. The final concentration of SARS-CoV-2 RNA in each sample ranged from 0.8 – 820 c/μL and the final concentration of human RNA was held constant across all samples at 2000 c/μL. The same conditions were tested for both the CDC singleplex test and the multiplex test. Thermal cycling conditions for each MM are outlined in **Table 1**.

## Results

### In silico analysis

In silico analysis predicted unfavorable interactions were unlikely between N1, N2, and RP primers and probes, even if primers and probes were subjected to lower temperature and higher oligo concentration assay conditions. These results suggested that primers and probes from the singleplex reactions could be combined into a multiplex reaction without substantial loss in sensitivity and minimal risk of spurious product formation.

### Developing a multiplex RT-PCR for SARS-CoV-2

For this work, purified SARS-CoV-2 RNA was used to evaluate the performance of a multiplex RT-PCR, consisting of N1, N2, and RP targets from the CDC singleplex test. At a 1:1 ratio of SARS-CoV-2 RNA to human RNA, all targets were detectable down to 4 c/µL of sample of each RNA per reaction, **Figure 1A**. The SARS-CoV-2 input was also held at 0.8, 2.0, 4.0, or 8.0 c/µL of sample while human RNA input was increased from 10^1^ – 10^4^ c/µL of sample. Both SARS-CoV-2 targets (N1 and N2) were detectable for all replicates of 4 c/µL of sample regardless of the amount of human RNA present, **Figure 1B-D**. When SARS-CoV-2 RNA was present at 2 c/µL of sample with higher amounts of human RNA (>10^3^ c/µL) there were dropouts of either N1 or N2 replicates suggesting slight competitive inhibition. At very low concentrations of SARS-CoV-2 RNA (0.8 c/µL), the N1 and N2 targets performed in a stochastic manner.

**Figure 1.**
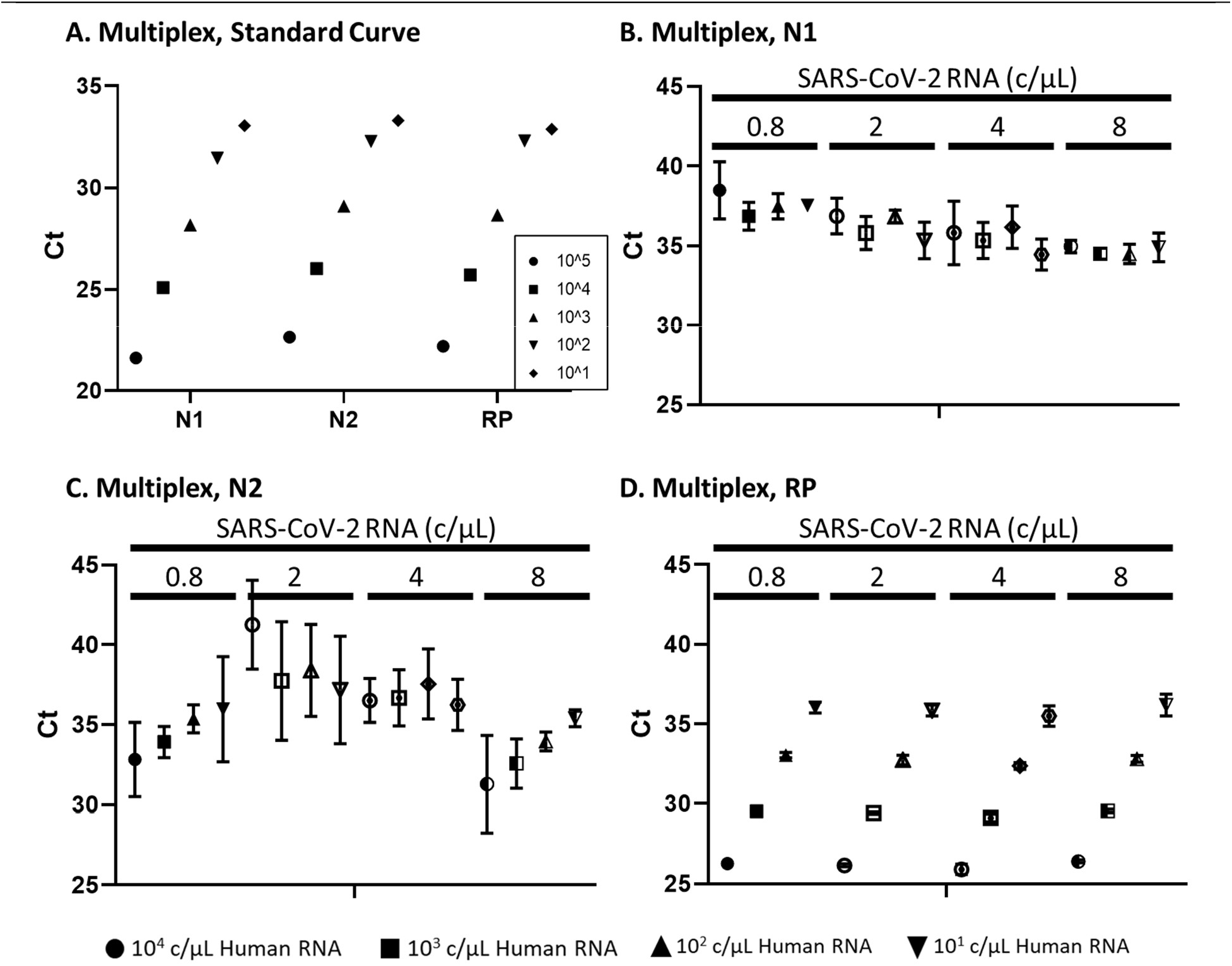
Comparing the CDC singleplex test and multiplex reaction for detection of SARS-CoV-2 and human RNA using the TaqPath MM (ThermoFisher). **A**. When the SARS-CoV-2 and human RNA ratio is held constant at 1:1, all targets are detectable down to 4 c/µL sample of each RNA. Detection of N1 (**B**.), N2 (**C**.), and RP (**D**.) in the multiplex reaction with varying concentrations of human RNA. At the lower end of SARS-CoV-2 RNA input, all targets were detectable at 4 c/µL of sample when human RNA ranged from 10^1^ – 10^4^ c/µL of sample.

We also tested the specificity of the multiplex RT-PCR using 78 clinical samples from Discovery Life Sciences. All samples were SARS-CoV-2 negative and 17 were positive for a variety of other respiratory infections including influenza A and B, respiratory syncytial virus, pneumoniae, and coronavirus NL63, HKU1, or OC43. None of these samples were positive for SARS-CoV-2 when tested with the multiplex reaction, **Table S2**.

After validating the analytical performance of the multiplex assay, we compared singleplex and multiplex detection of SARS-CoV-2 RNA purified from 90 patient samples. We observed 86% concordance with SARS-CoV-2 positive samples and 98% concordance with negative samples, **Table 2**. While many of the results were concordant, we observed the N2 signal in the multiplex reaction dropped below the established LoD more frequently than in the original, CDC singleplex test. To identify a sample as positive, the CDC EUA requires both N1 and N2 to be detected. For all the discordant positive samples, N1 was positive while N2 was negative leading to an overall negative result. Any Ct above 40 was considered a negative result to align with MIQE guidelines(21). The full set of data for each clinical sample is provided in **Table S3**.

**Table 2.**
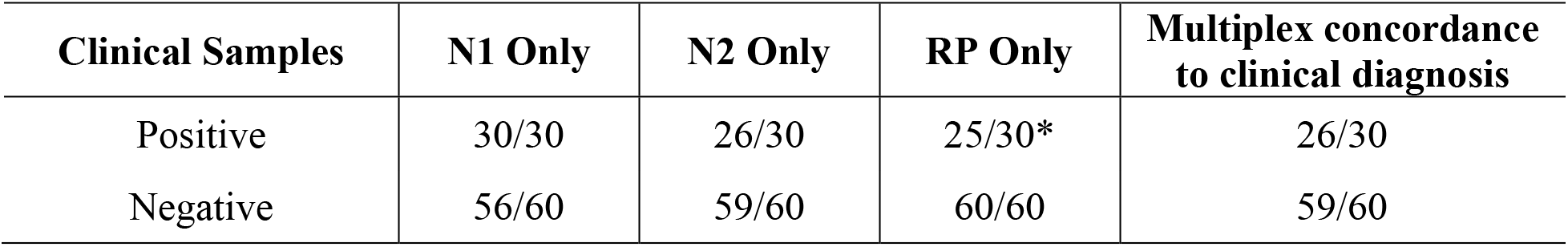
Multiplex RT-PCR concordance to the CDC singleplex test using RNA purified from 90 clinical NP samples. The data are provided as individual targets as well as overall sample determination. Note, for these data a sample is considered positive if both N1 and N2 are detected based on the CDC EUA recommendation. *All five SARS-CoV-2 positive samples with no RP signal had very high levels of SARS-CoV-2 RNA (>10^5^ c/µL), suggesting that the pathogen targets competitively inhibit the human target during amplification. Ct values and viral loads for each sample are provided in **Table S3**.

| Clinical Samples | N1 Only | N2 Only | RP Only | Multiplex concordance to clinical diagnosis |
| --- | --- | --- | --- | --- |
| Positive | 30/30 | 26/30 | 25/30* | 26/30 |
| Negative | 56/60 | 59/60 | 60/60 | 59/60 |

### Impact of viral transport media on RT-PCR

For this work, three common VTMs were tested in five commercially available RT-PCR MMs to determine the impact of VTM on SARS-CoV-2 detection. Purified SARS-CoV-2 RNA and human RNA were spiked into VTM or VTM diluted in 10 mM Tris at pH 8 (dilutions tested 1:10, 1:100, and 1:1000). The fold-change in Ct values was calculated to determine the impact on amplification (see the Supporting Information for more details). If a condition did not have a Ct value, it was set to 45, the maximum number of cycles, to determine a ΔCt.

The first set of experiments tested all VTM/MM combinations for each singleplex reaction: N1, N2, and RP, **Figure 2**. **Figure S1** reports the ΔCt for each stock VTM/MM combination. The TaqPath MM was the least inhibitor-tolerant across all three VTMs and at least one replicate had no detected Ct value for one or more of the targets indicating complete amplification inhibition. Both the UltraPlex and qScript MMs were the most tolerant and even showed some amplification enhancement in two of the three VTMs. The M4RT VTM resulted in the most amplification inhibition, significantly impacting three of the five MMs. Amplification enhancement and/or inhibition was reduced or entirely removed when VTM was diluted in 10 mM Tris (**Figure S2**).

**Figure 2.**
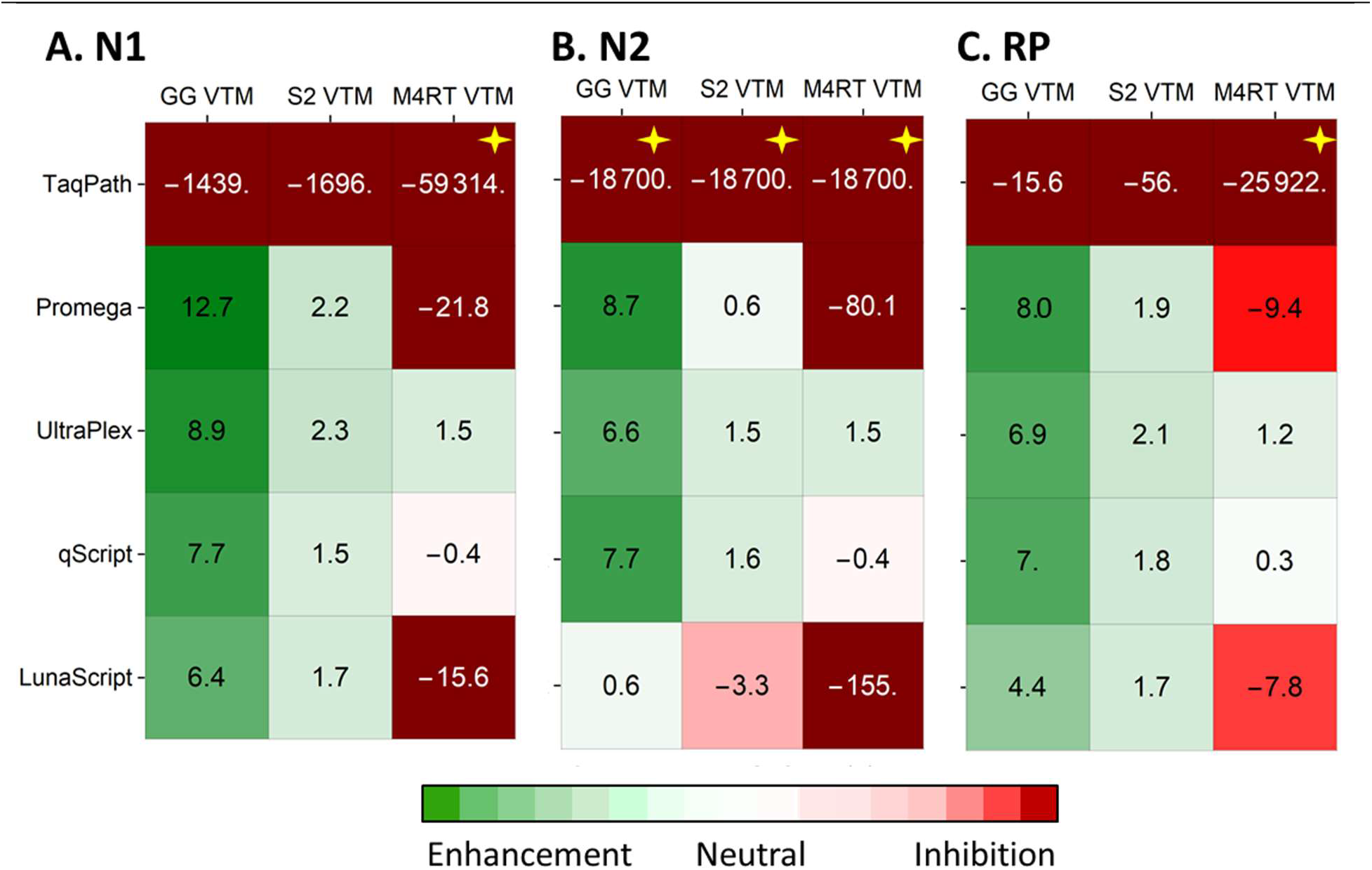
Impact of stock VTM on the CDC singleplex test. These data indicate that the TaqPath MM is the least inhibitor-tolerant while both the UltraPlex and qScript MMs were the most tolerant and even showed some amplification enhancement. Amplification enhancement and/or inhibition was reduced or entirely removed when diluted VTM was tested (**Figure S2**). The data presented are fold-change in Ct values between RNA spiked in VTM and control reactions of RNA spiked in 10 mM Tris buffer, pH 8. If a condition did not have a Ct value it was set to 45 to generate a difference in Ct. Conditions marked with a “✧" of the replicates did not have a Ct value after 45 cycles resulting in complete amplification inhibition. **Figure S1** reports the ΔCt for each condition. All reported values are averages of N=3 per condition. The SARS-CoV-2 and human RNA were kept constant at 820 c/µL and 2000 c/µL, respectively. **A**. Impact on SARS-CoV-2 N1 target, **B**. N2 target, and human RP target.

Next, we assessed the impact of VTM containing low viral RNA copies in the CDC singleplex test using the qScript MM, which was one of the most inhibitor tolerant MMs tested. For this work, SARS-CoV-2 RNA varied between 0.4 – 820 c/µL of sample, while the human RNA was kept constant at 2000 c/µL of sample.

VTM samples amplified in the CDC singleplex test using the qScript MM were able to reliably detect as few as 2 c/µL of sample, **Figure 3**. At lower inputs (<0.8 c/µL), amplification was not observed for all replicates of either the Tris control or VTM samples. In a few cases, the VTM samples resulted in amplification when the Tris controls did not, again suggesting amplification enhancement in the presence of some transport media.

**Figure 3.**
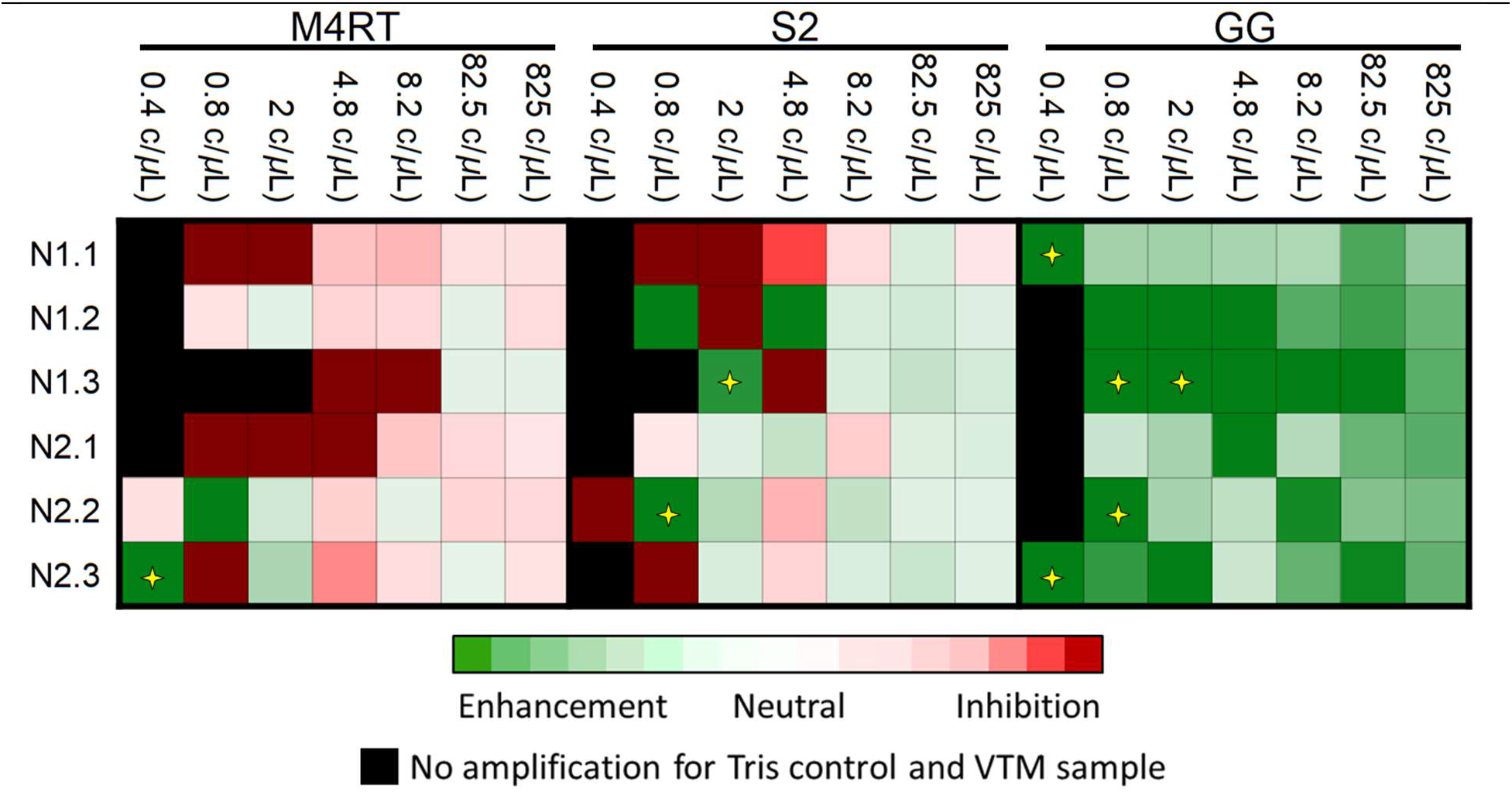
Impact of VTM on the CDC singleplex test using the qScript MM with low input copies of SARS-CoV-2 RNA. The data presented are fold-change in Ct values between RNA spiked in VTM and control reactions of RNA spiked in 10 mM Tris buffer, pH 8. Three replicates are shown for each condition. The black boxes indicate samples where no amplification signal was detected for both the Tris control and VTM sample. Conditions marked with a “✧” represent signal in the VTM sample, but no signal in the Tris control indicating potential amplification enhancement. VTM samples were able to reliably detect as little as 2 c/µL of sample.

From the clinical samples we received, the SARS-CoV-2 positives had viral loads that ranged between 1×10^0^ – 1×10^7^ c/µL with an average of 6×10^5^ c/µL (**Table S3**). These clinical viral load data with the low detection limit of the VTM in RT-PCR, **Figure 3**, suggest that amplification directly from clinical samples should accurately detect SARS-CoV-2 without purification.

In the presence of VTM, the multiplex reaction with qScript MM was able to detect SARS-CoV-2 RNA at 2 c/µL of sample for all replicates, regardless of the amount of human RNA, **Figure 4**. At lower inputs (<0.8 c/µL), amplification was not observed for all replicates of either the Tris control or VTM samples. In a few cases, the VTM samples resulted in amplification when the Tris controls did not, again suggesting amplification enhancement in the presence of some transport media. The same experiment was performed with the TaqPath MM, which showed significant amplification inhibition of across all VTMs tested (data not shown).

**Figure 4.**
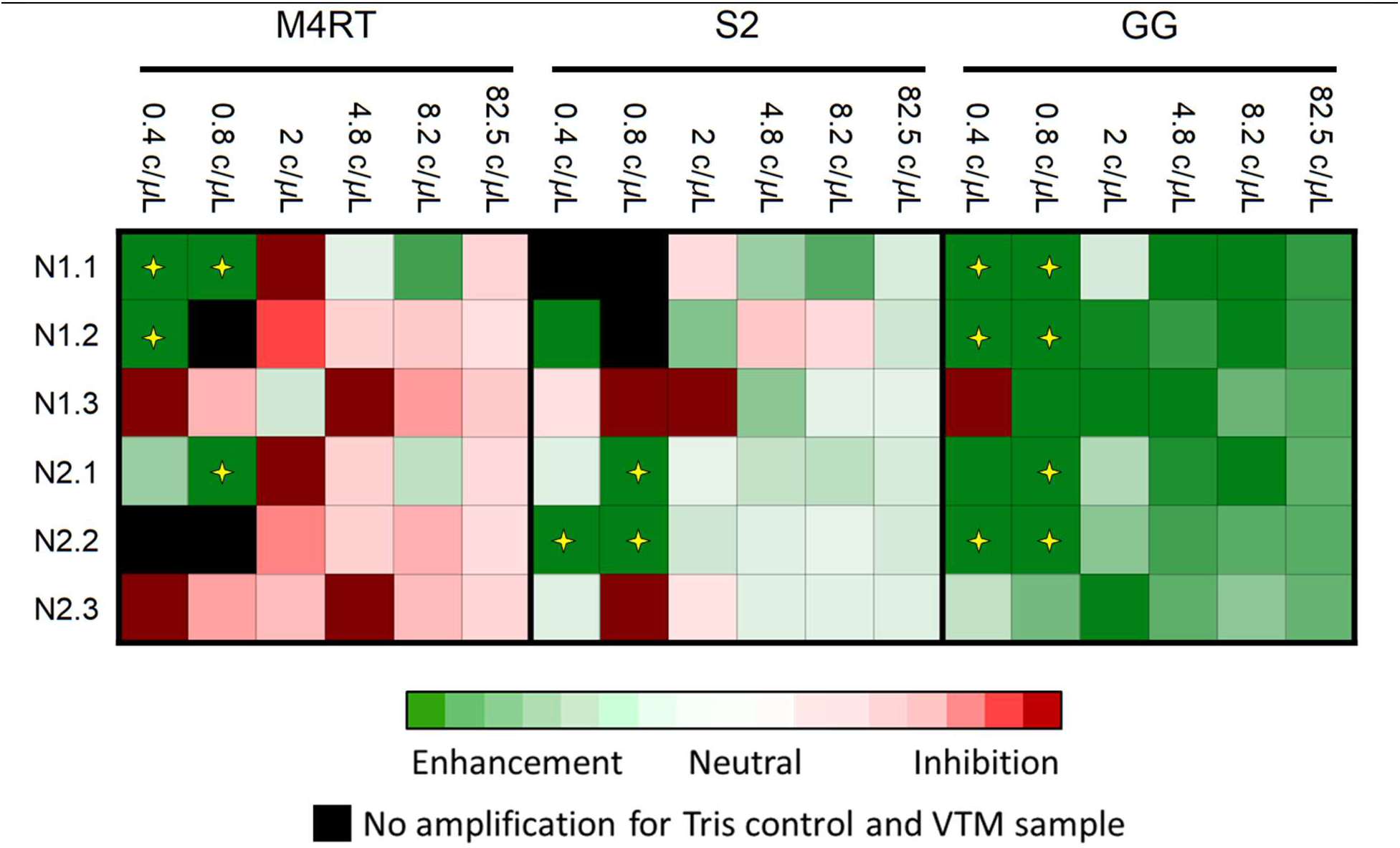
Impact of VTM on the multiplex test using the qScript MM with low input copies of SARS-CoV-2 RNA. The data presented are fold-change in Ct values between RNA spiked in VTM and control reactions of RNA spiked in 10 mM Tris buffer, pH 8. Three replicates are shown for each condition. The black boxes indicate samples where no amplification signal was detected for both the Tris control and VTM sample. Conditions marked with a “✧” represent signal in the VTM sample, but no signal in the Tris control sample indicating potential amplification enhancement. Direct multiplex amplification of VTM samples in the qScript MM was able to detect as few as 2 c/µL of sample for all replicates.

Next, we combined amplification directly from clinical samples with our multiplex reaction. This simplified workflow reduced the time-to-result for each sample by at approximately 1.5 hours due to the removal of RNA purification prior to amplification. The clinical samples (NP swabs in VTM) were received from the Washington State Department of Health (DOH) Public Health Laboratories (Shoreline, WA, USA). We tested 16 samples (eight SARS-CoV-2 positives and eight SARS-CoV-2 negatives) with direct amplification using the multiplex RT-PCR and qScript MM. The viral loads of the eight positive samples ranged from 3.0×10^0^ – 2.1×10^6^ c/µL of VTM (based on purified RNA using the CDC singleplex test). This range was specifically selected to represent the broad clinical viral loads observed for SARS-CoV-2 infection. With direct amplification of clinical samples using the multiplex reaction, we saw 8/8 concordance of the negative samples across all targets. For the SARS-CoV-2 positive samples, only six of the eight were detected, **Table 3**, with viral loads ranging from 2.5×10^1^ – 2.1×10^6^ c/µL of sample. Each sample was run with five replicates to determine concordance with the clinical diagnosis.

**Table 3.** Concordance of SARS-CoV-2 diagnosis to detection results using our multiplex RT-PCR with direct amplification from 16 clinical NP samples in VTM. These data were generated with the qScript MM. The provided data are separated by results from individual targets as well as the final concordance call (Yes or No). All samples were run five times and the average result was used to determine concordance. Note, for these data, a sample is considered positive if both N1 and N2 is positive, as recommended by the CDC(5). Additionally, any Ct above 40 was considered negative in alignment with standard practices(21).

| Patient Sample ID | SARS-CoV-2 (c/μL) | N1 Only | N2 Only | RP Only | Multiplex concordance to clinical diagnosis |
| --- | --- | --- | --- | --- | --- |
| **SARS-CoV-2 Positive Samples** |  |  |  |  |  |
| WA0364736 | 2.1x10<sup>6</sup> | 5/5 | 5/5 | 5/5 | Yes |
| WA0364714 | 5.0x10<sup>5</sup> | 5/5 | 5/5 | 5/5 | Yes |
| WA0364715 | 6.3x10<sup>4</sup> | 5/5 | 5/5 | 5/5 | Yes |
| WA0364723 | 1.4x10<sup>4</sup> | 5/5 | 5/5 | 5/5 | Yes |
| WA0364702 | 8.3x10<sup>3</sup> | 5/5 | 5/5 | 5/5 | Yes |
| WA0364405 | 2.7x10<sup>2</sup> | 0/5 | 0/5 | 5/5 | No |
| WA0364810 | 2.5x10<sup>1</sup> | 3/5 | 4/5 | 5/5 | Yes |
| WA0364834 | 3.1x10<sup>0</sup> | 1/5 | 2/5 | 5/5 | No |
| **SARS-CoV-2 Negative Samples** |  |  |  |  |  |
| WA0361635 | - | 5/5 | 5/5 | 5/5 | Yes |
| WA0361636 | - | 5/5 | 5/5 | 5/5 | Yes |
| WA0361637 | - | 5/5 | 4/5 | 5/5 | Yes |
| WA0361638 | - | 5/5 | 5/5 | 5/5 | Yes |
| WA0361639 | - | 5/5 | 4/5 | 5/5 | Yes |
| WA0361640 | - | 5/5 | 5/5 | 5/5 | Yes |
| WA0361641 | - | 5/5 | 4/5 | 5/5 | Yes |
| WA0361642 | - | 5/5 | 4/5 | 5/5 | Yes |

## Discussion

### Developing a multiplex RT-PCR for SARS-CoV-2

We show that the CDC singleplex test and our multiplex reaction LODs were comparable at 1 c/µL and 4 c/µL, respectively, using the TaqPath MM. We were able to achieve a slightly improved LoD of 2 c/µL for the multiplex test when using the qScript MM and RNA spiked into VTM. Additionally, the multiplex reaction tested with purified RNA from 90 clinical samples showed 86-98% concordance with clinical diagnoses for positive/negative SARS-CoV-2 results, **Table 2**. Up to three replicates were run for these samples to determine concordance, **Table S4**.

For the multiplex reaction, we observed unexpected results for the N2 target when increasing the amount of human RNA in the reaction. When the N2 concentration is held constant, the Ct appears to decrease, rather than remain constant, as the human RNA concentration increases. We believe this to be signal carryover from the human RP target in the Cy5 channel, **Figure 1C**.

In **Table 2**, all the discordant positive samples for the multiplex test detected the N1 target, but not the N2 target. We reported a concordant positive only if both N1 and N2 were positive to align with the recommendations from the CDC singleplex test(5). Currently, there are two commercial assays that only require a single SARS-CoV-2 target for determining a positive sample(22,23). If our data used these guidelines, the positive concordance would be 100%. However, using only N1 to determine a positive result would also impact our negative concordance and drop it from 98% to 93% because there would be three additional false positive samples detecting N1 and not N2. For the SARS-CoV-2 negative samples, 40 cycles was used as the cut off value to align with the MIQE guidelines(21).

### Impact of viral transport media on RT-PCR

Most clinical samples collected for the current COVID-19 outbreak are stored in VTM before they are tested. There are multiple variations of VTM and UTM which maintain the quality of clinical specimens during collection, transport, and long-term freezer storage because few diagnostic tests are authorized to run at the point of sample collection. With this well-established clinical practice in mind, ideal COVID-19 tests would be tolerant of VTM to reduce the need for sample clean-up prior to testing. Additionally, because a consistent supply of a single VTM may be unreliable during this pandemic, optimal diagnostic tests would be tolerant of a range of transport media. Based on the results above in Figure 2, both the qScript and UltraPlex perform well in combination with all VTMs and with low SARS-CoV-2 viral loads. The data from mock samples of RNA spiked into VTM suggests that RNA purification is not necessary for COVID-19 detection.

Finally, we tested extraction-free amplification of SARS-CoV-2 from patient samples using the multiplex RT-PCR. These results were promising with 14/16 clinical samples correctly identified. On-going work is underway to explore additional methods to improve detection direct from VTM samples including the addition of a simple heat-lysis step prior to amplification. Previous reports have suggested a heat step may increase the accessibility of the viral RNA therefore improving amplification efficiency(24).

Recently, there have been reports of up to 33% false negative results with FDA authorized systems such as the Abbott ID NOW(25). One of the most attractive features of this test is the simplified workflow: NP samples added directly to the test. Our data also demonstrate a simplified workflow by directly amplifying samples from NP swabs, and it enables more flexibility in reagent selection and shows increased specificity.

Overall, this work suggests that eliminating up-front RNA purification and simplifying the RT-PCR amplification from three singleplex reactions to a multiplex test can significantly reduce complexity, time, and costs for detecting COVID-19 with the appropriate selection of MM. On-going work will explore additional pathways to further streamline this simplified workflow and expand testing to larger numbers of clinical samples to further validate the approach.

### Supporting Information

Supporting Information is available online. The text includes two figures, four tables, and some additional methods.

## Data Availability

All data is provided in the Supporting Information.

## Acknowledgments

Funding was provided by the Bill and Melinda Gates Foundation Trust through The Global Good Fund I, LLC (www.globalgood.com) at Intellectual Ventures. The Global Good Fund I, LLC provided support in the form of salaries for SAB, RG, AS, CB, RR, CO, STM, PJ, BHW, and JTC, as well as financial support for the study, but did not have any additional role in study design, data collection and interpretation, decision to publish, or preparation of the manuscript.

We also thank the Washington State DOH Public Health Laboratories for providing clinical samples and Joshua Bishop from Intellectual Ventures Laboratory for valuable discussions.

## Notes

### Competing Interest Statement

The authors have declared no competing interest.

### Author Declarations

All clinical samples used in this study were discarded, de-identified samples and therefore did not require IRB approval. We've updated the manuscript to reflect this information.

